# Serum uric acid and total bilirubin as putative biomarkers of resistance in Prodromal Parkinson’s disease: Longitudinal data from the PPMI study

**DOI:** 10.1101/2021.12.04.21267290

**Authors:** Christos Koros, Athina-Maria Simitsi, Anastasia Bougea, Nikolaos Papagiannakis, Andreas Prentakis, Dimitra Papadimitriou, Ioanna Pachi, Efthalia Angelopoulou, Ion Beratis, Efthymia Efthymiopoulou, Konstantinos Lourentzos, Maria Bozi, Sokratis G. Papageorgiou, Xenia Geronicola Trapali, Anastasios Bonakis, Maria Stamelou, Leonidas Stefanis

## Abstract

**Background:** The role of blood uric acid and more recently bilirubin as biomarkers in symptomatic motor PD has been increasingly established in the literature.

**Objective:** Our present study assessed the role of serum uric acid and total bilirubin as putative biomarkers in a prodromal PD cohort followed longitudinally.

**Methods:** Longitudinal 5-year serum uric acid and total bilirubin measurement data of 65 Prodromal PD patients (including REM Sleep Behavior disorder (RBD), N=39 and Hyposmia, N=26) with an abnormal DATSCAN imaging were downloaded from the Parkinson’s Progression Markers Initiative (PPMI) database. This cohort was compared with 423 de novo sporadic PD patients and 196 healthy controls enrolled in the same study.

**Results:** After adjusting for age, sex and Body Mass Index (BMI), baseline and 5-year longitudinal serum uric acid levels were higher in the Prodromal cohort and RBD subgroup as compared to the motor PD cohort. This was also true for longitudinal measurements in the Hyposmic subgroup. In contrast, baseline and longitudinal serum total bilirubin did not differ between each prodromal group and the PD cohort.

**Conclusions:** Our results are indicative of a role of serum uric acid (but probably not of total bilirubin) as a marker of neuroprotection, in a certain subgroup of premotor patients exhibiting exclusively non motor features (hyposmia or RBD). It is possible that an inherent antioxidant resistance of a subset of RBD or hyposmia patients with high serum uric acid level delayed or precluded the emergence of a motor PD phenotype as opposed to the PD cohort.

## Introduction

Prodromal Parkinson’s disease (PD) corresponds to the premotor phase of PD lasting many years. During this phase the neurodegeneration process is active and ongoing and typical prodromal non-motor symptoms like REM sleep behavior disorder (RBD), depression, hyposmia and constipation emerge. The progression to the nonmotor and motor manifestations of PD aligns well with the neuropathological staging system proposed by Braak [1]. Numerous risk factors have been thoroughly described in previous studies and they facilitate the assessment of the probability to develop motor PD [2]. Such markers have been incorporated in the recent MDS criteria for prodromal PD [3].

Oxidative stress is thought to play a vital role in the pathogenesis of Parkinson’s disease (PD)[4]. Two endogenous antioxidant molecules, uric acid and bilirubin, are considered to be buffering agents and to likely protect dopaminergic neurons from oxidative stress in PD. Uric acid exerts its antioxidant function mainly by means of ferrum chelation, although other mechanisms may also contribute to this effect. In vitro studies have shown that uric acid could induce autophagy activation and ameliorate alpha-synuclein (SNCA) accumulation [5]. In an in vivo PD model, uric acid demonstrated neuroprotective properties for dopaminergic neurons by means of increased expression of nuclear factor E2-related factor 2 (Nrf2) and three Nrf2-responsive genes [6]. In certain PD population studies, PD patients exhibited lower serum uric level in comparison to healthy controls [7,8] but in other cohorts this difference was less evident [9]. As far as prodromal PD is concerned, large retrospective and prospective studies have demonstrated decreased serum uric acid in individuals that eventually developed motor PD during follow up [10]. High serum uric acid levels have been correlated with lower risk of PD, a milder disease course and with the more benign tremor dominant (TD) motor subtype [11,12]. As far as cognitive outcomes are concerned, lower levels of serum uric acid in the early disease stages were reported to be associated with the later occurrence of mild cognitive impairment (MCI) in an early PD cohort [13]. However, the causal relationship between high plasma urate and low risk of PD and its putative prognostic value have been questioned by other researchers [14,15].

Increased serum bilirubin levels might also be related to PD etiopathology as a possible serum biomarker of oxidative stress dysregulation. The heme oxygenase (HO) - bilirubin pathway is one of the major antioxidant defense mechanisms. Heme oxygenase-1 (HO-1), an enzyme that converts heme to free iron, carbon monoxide (CO) and biliverdin (bilirubin precursor) is expressed in response to various stressors including neurodegenerative disorders like Parkinson’s disease [16]. Individuals with decreased serum bilirubin probably lack the endogenous defense system to prevent free radicals from damaging dopaminergic cells in the substantia nigra. Under control of different transcription factors but with a prominent role played by Nrf2 (common mediator also implicated in uric acid antioxidant pathway), HO-1 induction is crucial in nervous system response to damage [17]. In previous studies, PD subjects showed higher levels of bilirubin compared with controls, possibly due to HO overexpression as a compensatory mechanism from the early stages of PD; higher bilirubin levels in PD were not likely to be related to dopaminergic replacement therapy [18,19]. Lee and co-authors showed using imaging correlates, that bilirubin was the most significant antioxidant marker and it was correlated to the degree of dopaminergic deficit [20].

The Parkinson’s progression markers initiative (PPMI) study evaluated the 5-year change of clinical, imaging and biochemical parameters in de novo PD patients, in subjects with prodromal PD symptoms (RBD and hyposmia) and in healthy controls. The aim of our present study is to determine whether there are baseline and longitudinal differences in the aforementioned biomarkers, serum uric acid and total bilirubin level between Prodromal individuals and Parkinson’s disease (PD) patients or healthy controls enrolled in the PPMI study.

## Materials & Methods

Data used in the preparation of this article were obtained from the Parkinson’s Progression Markers Initiative (PPMI) database (www.ppmi-info.org/data). For up-to-date information on the study, visit www.ppmi-info.org. The present study was conducted in agreement with the principles of the Declaration of Helsinki. Signed informed consent was obtained from all participants recruited. The study was approved by the Scientific Board of all PPMI sites involved (including the Scientific Board of Attikon and Eginition hospitals).

Longitudinal 5-year serum uric acid and total bilirubin measurement data of 65 Prodromal participants were downloaded from the PPMI database. Moreover, we obtained data from 423 de novo sporadic PD patients and 196 healthy controls from the same database.

The Prodromal cohort in the PPMI study included premotor subjects with prodromal PD symptoms (REM sleep behavior disorder-RBD or hyposmia/reduced olfaction) [21,22]. RBD was diagnosed based on clinical history and sleep related scales along with polysomnography, if available. In the hyposmia cohort, olfaction was measured using the University of Pennsylvania Smell Identification Test (UPSIT).

Hyposmia was defined as a score of <10th percentile for age and sex. PPMI inclusion criteria enriched the RBD and hyposmia cohorts, with individuals with dopamine transporter binding deficit. Most participants with a normal DATSCAN were excluded from the study with the exception of 10% of those without a DAT binding deficit with the goal of keeping site investigators blinded to DAT SPECT results. A total number of 65 Prodromal subjects (RBD, n = 39 and Hyposmic, n=26) were finally enrolled. An additional group of 57 RBD and 93 Hyposmic individuals with a normal DATSCAN during the screening visit were excluded from follow up in PPMI (however serum uric acid and total bilirubin measurements from samples of this cohort collected during the screening visit were also available in the database of the study).

Biochemical analyses (including measurements of uric acid and total bilirubin level in serum) have been carried out in Covance laboratories in a uniform fashion, as per the study protocol.

Statistical analysis for baseline comparisons between the prodromal subjects (Prodromal cohort, RBD and Hyposmic subgroups) and the sporadic PD cohort / healthy controls was performed using univariate ANCOVA. Factors that could have an impact on uric acid [age/sex/ Body Mass Index (BMI)] or total bilirubin level (age/sex) were used as covariates in the analysis. Repeated measures ANCOVA (tests of within- and between-subjects effects) was used to examine the 5-year longitudinal changes of uric acid or total bilirubin level (Prodromal cohort, RBD and Hyposmic subgroups vs sporadic PD and healthy controls). Statistical significance was set at p< 0.05.

Pearson or Spearman correlations (according to the normality of group values) were calculated between uric acid level, duration since initiation of RBD and various baseline parameters (MDS-UPDRS score part III, 2-year and 4-year change in MDS UPDRS III and Montreal Cognitive Assessment MoCa score) in the RBD cohort, the hyposmia cohort and the prodromal cohort.

Finally, in order to assess the putative impact of uric acid levels on clinical progression of patients in the prodromal cohort and determine if high and low uric acid groups follow different trajectories, we compared high serum uric acid level- and low serum uric acid level-subgroups (at screening) in terms of motor deterioration (UPDRS III score at baseline, UPDRS III score on year 2, on year 4 and the 2-year and 4-year score change) using univariate ANCOVA with age, sex and BMI as covariates. The statistical analyses were performed using commercially available software (SPSS, Version 20.0).

## Results

### 3.1 Uric acid

After adjusting for age, sex and BMI, baseline serum uric acid measurements were higher in the prodromal cohort as compared to the PD cohort (p=0.040). 5-year longitudinal serum uric acid measurements were also higher in the Prodromal cohort vs PD (Between subjects effects, p=0.003). Moreover, there was no significant effect of Time but a significant effect of Time*Group interaction on uric acid level (Within subject effects, p= 0.050 and p=0.042 respectively). Baseline and longitudinal serum uric acid measurements did not differ statistically between Prodromal subjects and healthy controls (despite a trend for higher uric acid in the prodromal group) (p=0.487 and p=0.434). A significant effect of Time*Group interaction on uric acid level was evidenced (Within subject effects, p= 0.003) (Figure 1A).

**Figure 1.**
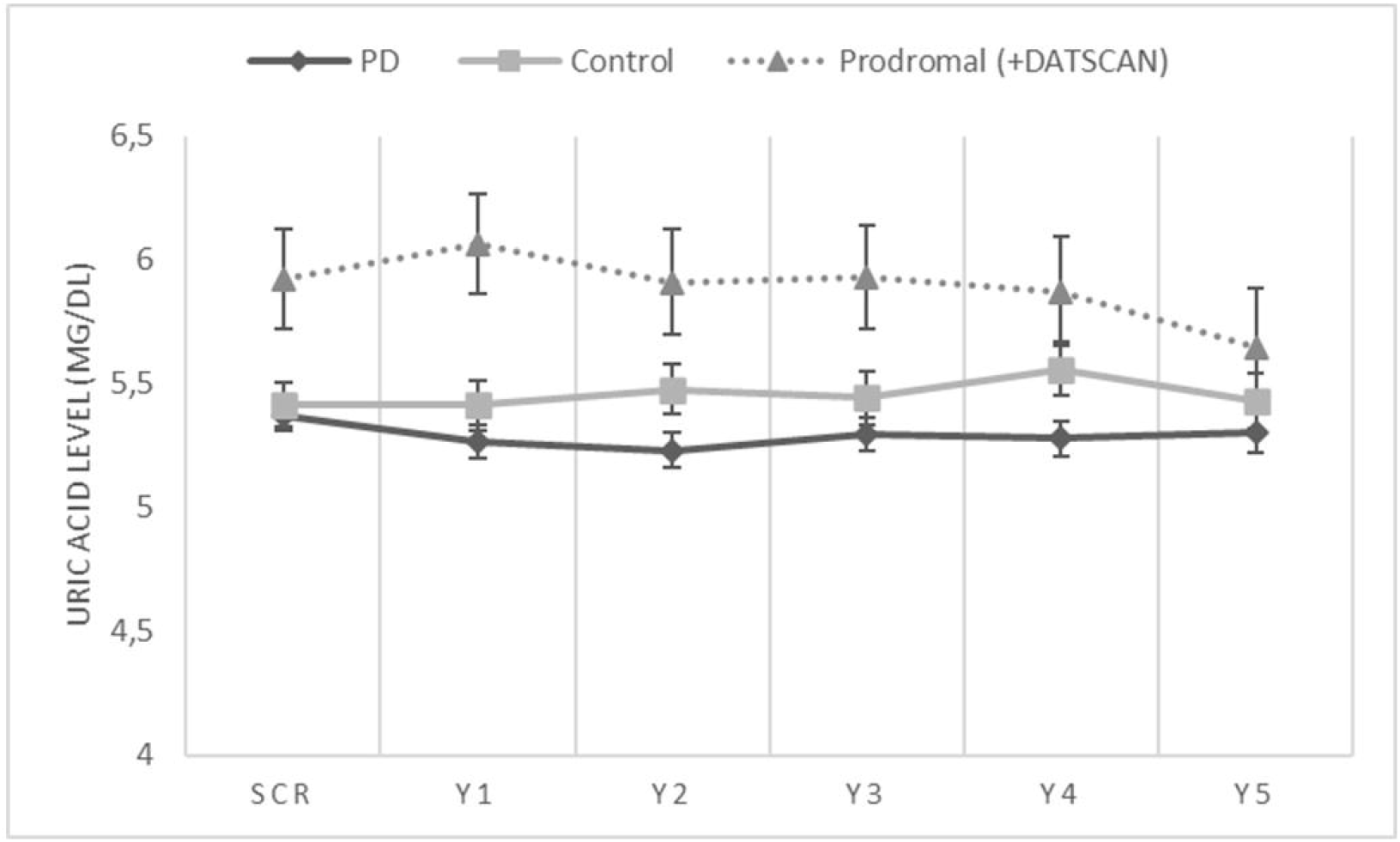
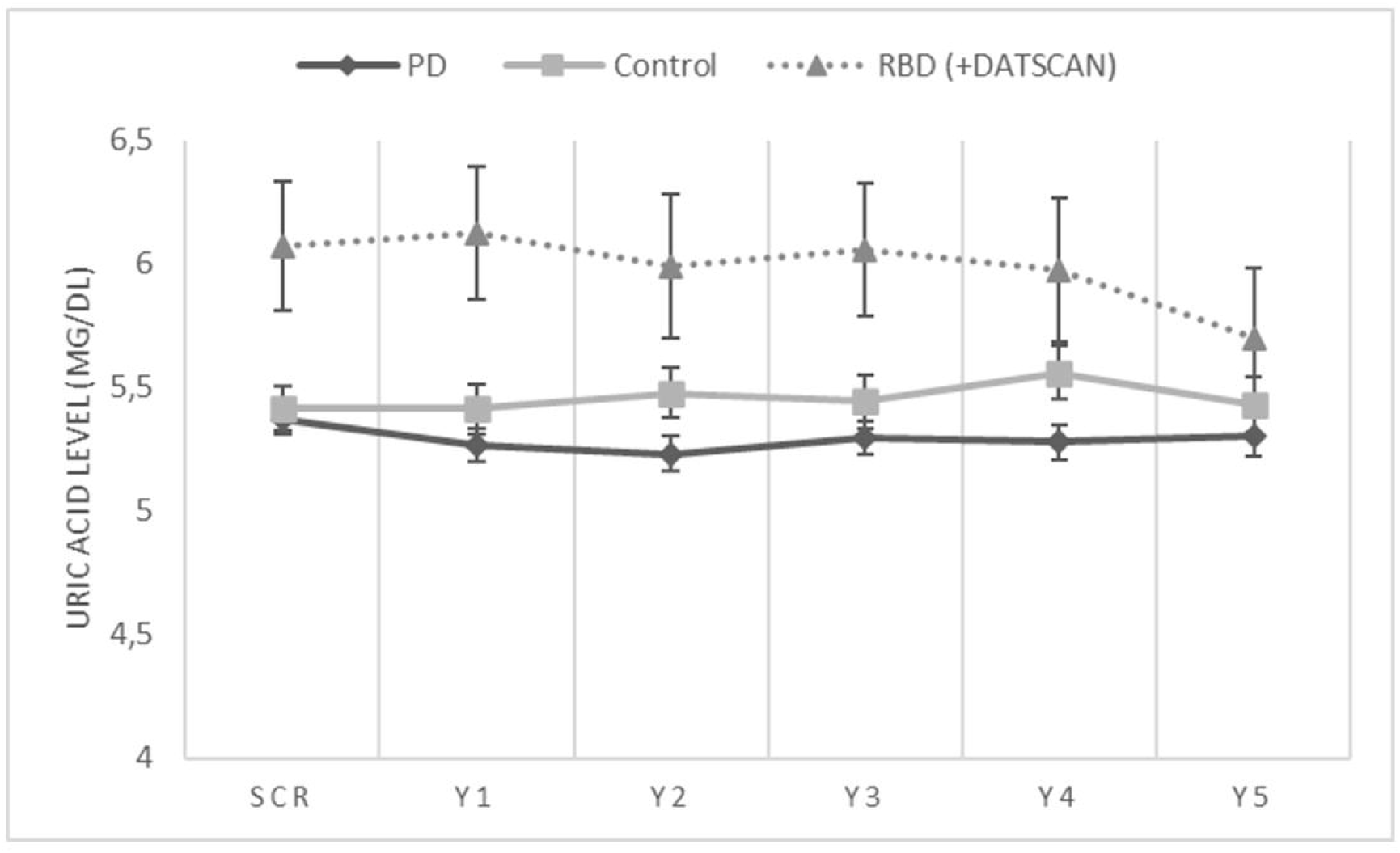
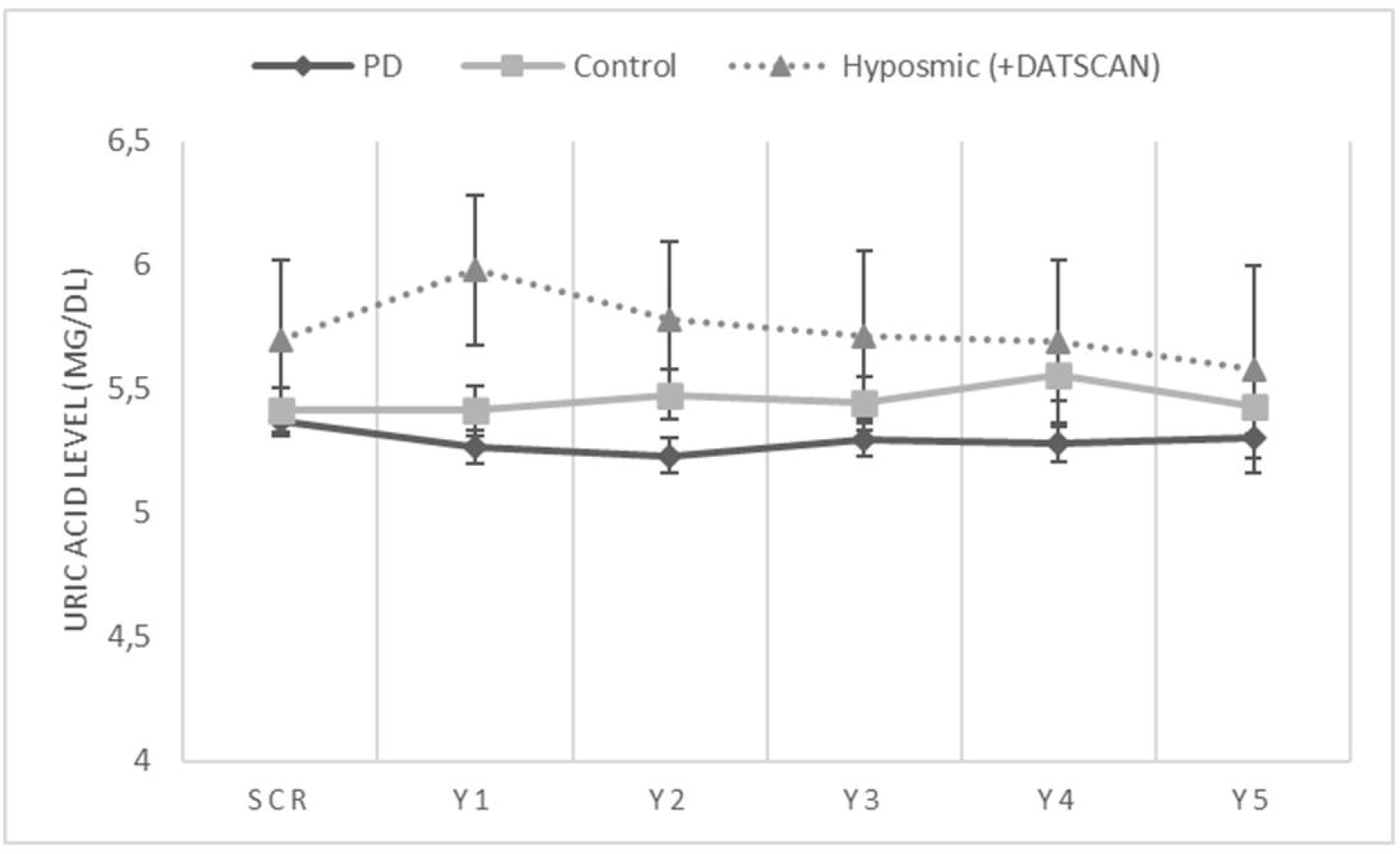
Longitudinal measurements of serum uric acid levels in: 1A) the Prodromal cohort vs sporadic PD and healthy controls, 1B) the RBD cohort vs sporadic PD and healthy controls, and 1C) the Hyposmic cohort vs sporadic PD and healthy controls. Statistical analysis has been performed using repeated measures linear ANCOVA (with age, sex and BMI as covariates). PPMI timepoints of visits SCR, V04, V06, V08, V10 and V12 correspond to Years 0, 1,2,3, 4 and 5 respectively. Error bars represent Standard Error of Mean.

Regarding each prodromal subgroup, baseline and 5-year longitudinal serum uric acid measurements were higher in the RBD cohort as compared to the PD cohort (p=0.028 and p=0.017 respectively). There was a significant effect of Time*Group interaction on uric acid level (Within subject effects, p= 0.043). Baseline and longitudinal serum uric acid measurements did not differ statistically between RBD subjects and healthy controls (despite a trend for higher uric acid in the RBD group) (Between subjects effects, p=0.279 and p=0.497). A significant effect of Time*Group interaction on uric acid level was evidenced (Within subject effects, p= 0.004) (Figure 1B). Furthermore, longitudinal (Between subjects effects, p=0.019) but not baseline serum uric acid measurements (p=0.354) were higher in the Hyposmic cohort as compared to the PD cohort. There was no significant effect of Time or Time*Group interaction on uric acid level (Within subject effects, p= 0.081 and p=0.674)). Finally, baseline and longitudinal serum uric acid measurements did not differ statistically between Hyposmic subjects and healthy controls (despite a trend for higher uric acid in the Hyposmic group) (p=0.905 and p=0.388) and no significant effect of Time or Time*Group interaction on uric acid level was evidenced (Within subject effects, p= 0.833 and p=0.282) (Figure 1C).

In the RBD group, we observed a positive correlation between the duration of RBD (time since first reported RBD symptoms) and the increases in MDS-UPDRS score part III scores between baseline and year 2 [Spearman correlation: r= 0.467 (p=0.004)] and between baseline and year 4 [Spearman correlation: r= 0.381 (p=0.024)]. There was no significant correlation between baseline uric acid level or 2-year and 4-year change in uric acid and the duration of RBD symptoms. Furthermore, there was no significant correlation between baseline uric acid level or 2-year and 4-year change in uric acid and either yearly MDS-UPDRS III scores (bl to year 5) or the increases in MDS-UPDRS score part III scores between baseline-year 2 and between baseline-year 4. No other parameter including Moca score correlated to baseline uric acid level in a statistically significant way. Finally, we failed to reveal any significant impact of initial uric acid levels on the severity of motor deterioration when comparing high and low uric acid subgroups.

### 3.2 Total Bilirubin

After adjusting for age and sex, baseline and longitudinal serum total bilirubin measurements did not differ between the Prodromal cohort and PD cohort (p=0.995 and 0.323 respectively). Moreover, there was no significant effect of Time or Time*Group interaction on total bilirubin (Within subject effects, p= 0.271 and p=0.122 respectively). Baseline and longitudinal total bilirubin measurements did not differ statistically between Prodromal subjects and healthy controls either (p=0.385 and p=0.756). No significant effect of Time or Time*Group interaction on total bilirubin level was evidenced (Within subject effects, p=0.633 and p=0.424) (Figure 2).

**Figure 2.**
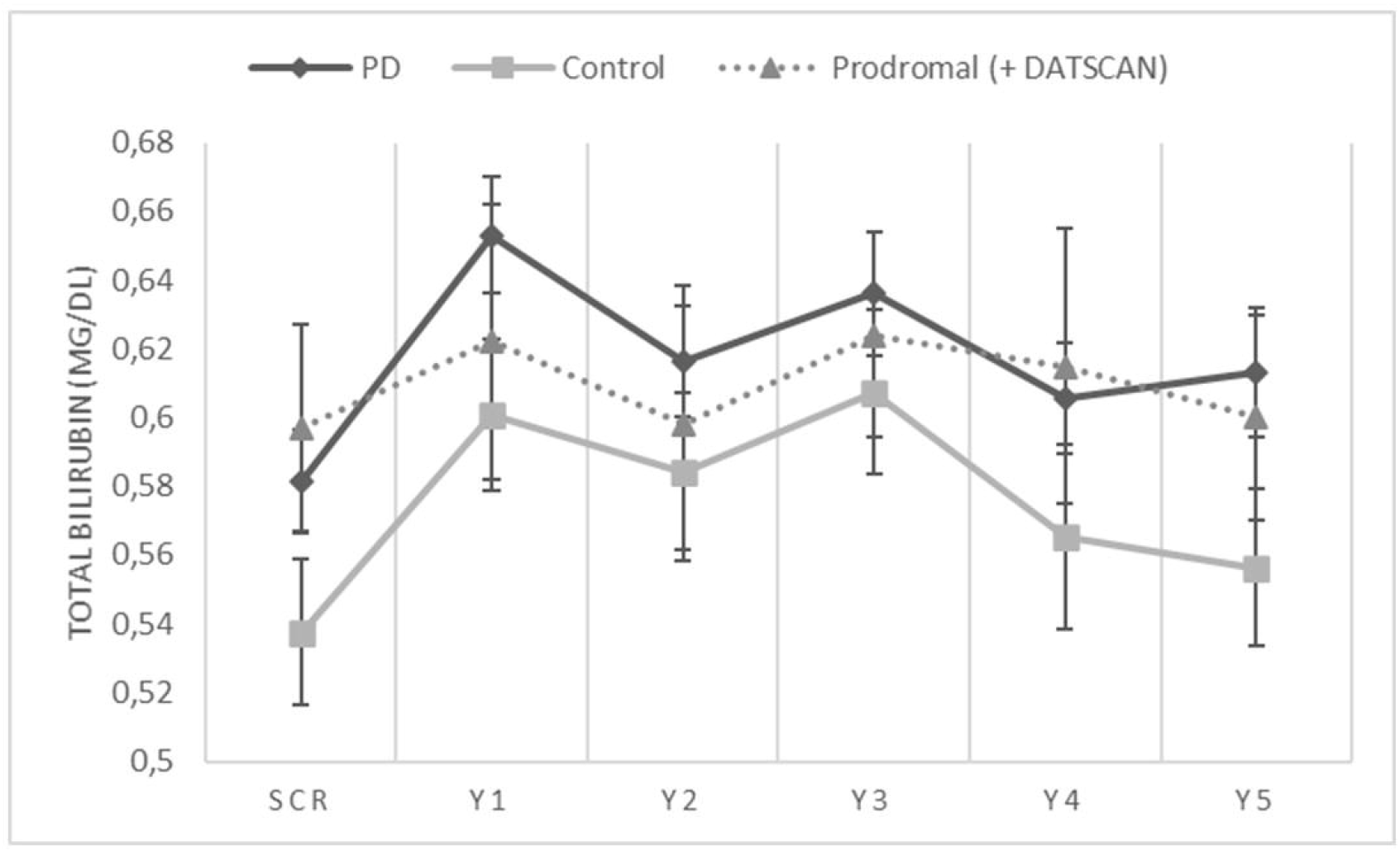
Longitudinal measurements of serum total bilirubin levels in the Prodromal cohort vs sporadic PD and healthy controls. Statistical analysis has been performed using repeated measures linear ANCOVA (with age and sex as covariates). PPMI timepoints of visits SCR, V04, V06, V08, V10 and V12 correspond to Years 0, 1,2,3, 4 and 5 respectively. Error bars represent Standard Error of Mean.

Similarly, baseline and longitudinal serum total bilirubin measurements did not differ between RBD and PD cohorts (p=0.348 and 0.219) and between RBD subjects and healthy controls either (p=0.84 and 0.644). Baseline and longitudinal serum total bilirubin measurements did not differ between Hyposmic cohort and PD cohort (p=0.264 and 0.872) and between hyposmic subjects and healthy controls (p=0.101 and 0.975 respectively). Moreover, there was no significant effect of Time or Time*Group interaction on total bilirubin in any of the subgroups.

## Discussion

The role of blood uric acid and more recently bilirubin as biomarkers in symptomatic motor PD has been increasingly established in the literature. Our present report assessed the role of serum uric acid and total bilirubin as putative biomarkers in a prodromal PD cohort followed longitudinally. It appears that both the entire Prodromal PD cohort and the RBD subgroup participants exhibited increased serum uric acid level as compared to sporadic PD both at baseline and during a 5-year follow up (this was also true for the hyposmic subgroups regarding follow up). The level of uric acid decreased longitudinally in the prodromal group, the RBD group and the Hyposmic group almost reaching that of sporadic PD at year 5. Moreover, despite a trend for higher uric acid, all three prodromal cohorts did not statistically differ from healthy controls following age, sex and BMI adjustments. Uric acid level increased in healthy controls as opposed to prodromal and PD subjects. On the other hand, total bilirubin did not differ between sporadic PD and either of the three prodromic cohorts (Prodromal, RBD and Hyposmic subgroups) and level of total bilirubin remained stable during the follow up period across the groups.

Our interpretation of these results focuses on the possible neuroprotective role of high serum uric acid, in a certain subgroup of premotor patients exhibiting exclusively non motor features (hyposmia or RBD). Such patients in the DATSCAN enriched PPMI prodromal cohort had clear-cut evidence of dopaminergic degeneration as exemplified by an abnormal DATSCAN imaging, however the presence of motor deficits might have been delayed due to an adequate endogenous antioxidant defense system. In contrast, symptomatic de novo PD patients might have proceeded earlier in their disease course to the motor phase of PD in part due to lessened anti-oxidant defenses. It is highly possible that an inherent resistance of some RBD or hyposmia patients delayed or precluded the emergence of a motor PD phenotype. During 5-year follow up a portion of RBD and Hyposmia patients converted to motor PD and some were started on dopaminergic treatment (13 out of 65 in the 5 year follow up). The gradual decrease in serum uric acid almost reaching the level of sporadic PD group reflects this situation (Figure 1A, B, C).

In accordance to our results, in a previous relevant study Uribe-San Martín and coauthors evaluated plasma urate levels in 24 patients with RBD and their role in the development of PD. Patients were divided into 2 groups according to the presence or absence of PD and it appeared that patients without PD and those who had more than 5 years of RBD exhibited higher levels of uric acid than patients with PD. The authors concluded that higher levels of plasma urate were associated with a longer duration of RBD without converting to PD, exhibiting resistance to the development of motor symptoms [23]. Moreover, in patients without PD, there was a positive correlation between years of evolution of RBD and the levels of uric acid. However, in our study, no such correlation between level of uric acid and duration of RBD symptomatology could be established in the RBD subgroup.

The notion of uric acid homeostasis contribution in the resistance of prodromal patients to PD could also apply to non manifesting carriers of pathogenic PD mutations. Bakshi and co authors evaluated the putative role of uric acid as a candidate biomarker of PD risk modulation in pathogenic LRRK2 mutation carriers using data from the LRRK2 Cohort Consortium and the Parkinson’s Progression Markers Initiative. According to their observations, non manifesting LRRK2 mutation carriers had significantly higher levels of uric acid than those who developed PD and this applied to both sexes. The authors reached the conclusion that uric acid monitoring could be used as a biomarker of resistance to PD among LRRK2 mutation carriers [24].

Regarding the association of uric acid level in prodromal PD with cognitive outcomes, no firm correlations could be established in our report. A previous study assessed how levels of serum uric acid affect cognition in patients with RBD. Regarding the memory domain, the low uric group had worse scores than the healthy controls, whereas the scores of high urate group were intermediate between the low uric acid group and the healthy controls. As far as other cognitive features like executive function and language domain are concerned, lower scores were found in the patients with low uric acid, whereas the scores of the patients with high urate were similar to those of the healthy controls.

According to the aforementioned study, uric acid levels affect cognitive function in patients with RBD, partly owing to its antioxidant and neuroprotective function [25]. In another study during the motor PD phase, uric acid and uric acid/creatinine levels in the early and medium stage PD patients were significantly higher than in the advanced stage ones [26]. Accordingly, if we consider prodromal and motor Parkinson’s disease to be the two edges in a continuum, higher uric acid could possibly mark the initial phases of the disorder and a gradual decrease of urate level might simply reflect disease progression.

The absence of significant differences in total bilirubin level between prodromal and motor PD might implicate a limited significance of this biomarker for the prodromal phase of the disorder. All current data on bilirubin focus on symptomatic PD patients with motor deficits [18,19]. Alternatively, since increased total bilirubin is a hallmark of early motor PD, it is possible that an adequate oxidative stress buffering during the prodromal phase of PD might hamper a rise in total bilirubin via the heme oxygenase 1 pathway. Clearly, further research is required in order to decipher the role of bilirubin as putative marker also for prodromal PD. In a study assessing the level of uric acid, uric acid/creatinine ratio, total bilirubin and indirect bilirubin, only bilirubin was associated negatively with PD stages. However, the lack of association between uric acid in the serum and total or indirect bilirubin suggests that these two biomarkers may play a different role in the etiopathogenesis of PD [27].

An important merit of our report is that data we used from the PPMI database have been collected and processed uniformly across PPMI centers. On the other hand, limitations include the relatively small number of Prodromal patients of the cohort studied in spite of a thorough clinical and laboratory assessment. Given the fact that clinical studies focused on uric acid related neuroprotective pharmacological treatments, like inosine administration, are underway in motor PD [28], the prodromal PD phase seems to be a promising field for early interventions as well. Future longitudinal studies targeted to elucidate the role of uric acid or bilirubin as putative biomarkers also for premotor PD might pave the way for relevant therapeutic interventions.

## Data Availability

All data produced in the present study are available upon reasonable request to the authors.

## Financial Disclosures of all authors (for the preceding 12 months)

Christos Koros received funding from the Michael J Fox Foundation for his participation in PPMI.

Athina-Maria Simitsi received funding from the Michael J Fox Foundation for her participation in PPMI.

Anastasia Bougea has no disclosures. Nikolaos Papagiannakis has no disclosures. Andreas Prentakis has no disclosures.

Dimitra Papadimitriou has no disclosures. Ioanna Pachi has no disclosures.

Efthalia Angelopoulou has no disclosures.

Efthymia Efthymiopoulou has no disclosures.

Konstantinos Lourentzos has no disclosures.

Ion Beratis received funding from the Micheal J Fox Foundation for his participation in PPMI.

Maria Bozi has no disclosures.

Sokratis G Papageorgiou has no disclosures.

Xenia Geronikola Trapali has no disclosures.

Anastasios Bonakis has no disclosures.

Maria Stamelou, serves on the editorial boards of Movement Disorders Journal and Frontiers in Movement Disorders, received travel and speaker honoraria from Abbvie, UCB and Specifar Pharmaceuticals, and the International Parkinson’s disease and Movement Disorders Society, receives research support from the Michael J Fox Foundation (PPMI), royalties from Oxford University Press, Cambridge University Press, and Elsevier.

Leonidas Stefanis is employed by the National and Kapodistrian University of Athens, Medical School and the Biomedical Research Foundation of the Academy of Athens. He has received the following grants : MULTISYN European Program (EU, FP7-HEALTH.2013.1.2-1, number 602646), PPMI (supported by the Michael J. Fox Foundation), IMPRIND-IMI2 Number 116060 (EU, H2020), SANTE Research Grant in Biomedical Sciences, NO-MND (EU-FP7-PEOPLE-2013-IRSES), NPF 2015 Investigator Award (Collaborator), “PBMC and urine collection in LRRK2 and idiopathic PD” Grant by the Michael J. Fox Foundation (Collaborator).

